# PERsonalised Knowledge to reduce the risk of Stroke (PERKS-International): a randomised controlled trial testing the efficacy of an mHealth application to reduce risk factors for the primary prevention of stroke

**DOI:** 10.64898/2026.03.19.26348870

**Authors:** Seana L Gall, Valery Feigin, Kate Chappell, Amanda G Thrift, Timothy J Kleinig, Dominique A. Cadilhac, Derrick Bennett, Mark R Nelson, Tara Purvis, Shabnam Jalili-Moghaddam, Gemma Kitsos, Rita Krishnamurthi

## Abstract

**Background and aims:** We evaluated the efficacy of the Stroke Riskometer™ mobile phone App to change the Life’s Simple 7® (LS7®) risk factor score at 6 months post-randomisation.

**Methods and design:** This Phase III, prospective, outcome assessor-blinded, 2-arm randomised controlled trial (RCT) in Australia and New Zealand recruited participants from August 2021 to January 2024. Inclusion criteria: age ≥35 and ≤75 years; ≥2 risk factors; smartphone ownership; no cardiovascular disease history. The intervention group was given access to the App; the usual care group received one e-mail with generic risk factor information. The primary outcome was the mean between group difference in LS7® (score 0 [poor] to 14 [ideal] comprising blood pressure, cholesterol, glucose, body mass index, smoking, physical activity and diet) from baseline to 6 months post-randomisation. Secondary outcomes were between group changes in individual LS7 items. Analyses were performed using intention to treat (ITT) principles with ANCOVA and linear mixed models to examine differences between groups, with pre-specified per protocol and subgroup analyses.

**Results:** We randomised 862 participants (mean ± SD age 58±11 years; 63% women; 74% Caucasian). At 6 months post-randomisation in ITT analyses, the mean difference between usual care (n=433) and intervention (n=429) groups in the change in LS7® score from baseline was 0.03 (95% CI -0.19, 0.25, p=0.79). Per protocol analyses (n=320 usual care; n=276 intervention) were similar (mean difference in change 0.11 95% CI -0.12, 0.34, p=0.34). Compared to usual care in ITT analyses, the intervention group had a borderline increase in metabolic equivalent of task (MET) minutes/week of physical activity (313.42 95% CI -2.80, 629.65, p=0.05), with no differences in other LS7® items.

**Discussion:** Among a general population aged 35 to 75 years with ≥2 stroke risk factors, there was no evidence that having access to the App changed overall LS7® scores at 6-month follow-up. Participants in the intervention group did have a small increase in physical activity, compared to the usual care group after 6 months, but not other individual risk factors.

## Introduction

Previously declining stroke incidence rates have plateaued, with some evidence of increasing incidence in younger age groups.^1^ It is estimated that up to 90% of strokes can be prevented because most are caused by risk factors that are modifiable through behaviour change or medications, for example high blood pressure, high cholesterol, diabetes and smoking.^2^ However, the prevalence of these risk factors across populations is very high. For example, in Australia, 23% of adults have high blood pressure, 66% are overweight or obese and 37% are physically inactive, with 30% have two or more risk factors.^3^ The prevalence of these risk factors is similar in other countries including New Zealand (40% physically inactive, 67% overweight or obese, 18% high blood pressure),^4^ the United States of America (48% high blood pressure,^5^ 40% obesity^6^) and the United Kingdom (30% high blood pressure, 64% overweight or obese).^7^ To prevent stroke, people need to be able to identify and manage their risk factors. Identifying interventions to prevent stroke, including strategies for the early identification and management of stroke risk, has recently been found to be a leading research priority.^8^

Interventions delivered by mobile phone, collectively known as mHealth interventions, are a potential solution to improve stroke prevention. mHealth leverages the fact almost 60% of the world’s population have access to smartphones.^9^ Emerging evidence suggests that mHealth apps may improve cardiovascular disease risk factors including lowering blood pressure,^10^ reducing body mass index (BMI)^11^ or increasing physical activity.^11^ However, there is a preponderance of small pilot studies, largely focused on people with pre-existing cardiovascular disease. There are few appropriately powered randomised controlled trials (RCTs) focused on primary prevention of cardiovascular diseases with objectively measured risk factors.^12^

The Stroke Riskometer™ is an mHealth application (herein termed ‘the App’) developed to help people identify and manage their stroke risk.^13^ It incorporates a validated algorithm including demographic, risk factor and health-related information to estimate a person’s absolute and relative risk of stroke within 5 and 10 years. The App incorporates evidence-based behaviour change tools including personalised feedback and information, self-monitoring of behaviour through tracking risk factors over time, credible sources of information and prompts through push notifications.^14^ In a pilot RCT the App was found to be feasible and acceptable with evidence of a small improvement in the Life’s Simple 7® (LS7®) behavioural and biomedical cardiovascular risk factor score^15^ after 6 months compared to a control group.^13^

The aim of this study was to examine the efficacy of the Stroke Riskometer App to improve the LS7® score from baseline to 6 months post-randomisation compared to a control group in people with two or more stroke risk factors.

## Methods

### Design

PERsonalised Knowledge to reduce the risk of Stroke (PERKS-International) was a Phase III, prospective, pragmatic, open-label, single-blinded end-point 2-arm RCT. Participants were recruited in Australia (Hobart, Tasmania; Melbourne, Victoria; Adelaide, South Australia) and New Zealand (Auckland, Hamilton). Mobile network coverage was high in all locations (>96% of population). The protocol for the study was published ^16^ and the trial was registered with the Australian and New Zealand Clinical Trials Registry (ACTRN12621000211864, date registered 1/03/2021).

The App has been available for free to download from the Apple App and Google Play stores since 2014. Therefore, to avoid contamination of the usual care group, participant recruitment materials did not refer specifically to the App. The trial was described as comparing two different ways of showing people their risk factors for stroke (see Supplementary Materials p16 for Participant Information Sheet and Consent Form). Support for users of the App was managed by the Auckland University of Technology. The trial design and implementation involved two people with lived experience of stroke. One was the chair of the Scientific Advisory Committee who provided oversight for the trial and the other was an investigator on the overarching grant (Synergies TO Prevent stroke [STOPstroke]) that funded this RCT.

The study is reported using the Consolidated Standards of Reporting Trials (CONSORT),^17^ the Template for Intervention Description and Replication (TiDIER) checklist^18^ and mHealth evidence reporting and assessment (mERA) guidelines (see Supplementary Material p3-10).^19^ All participants provided written informed consent. The study was approved by ethics committees in Tasmania (Tasmanian Health and Medical Human Research Ethics Committee H0023615); South Australia (Central Adelaide Local Health Network Research Governance 2021/SSA00461); Melbourne (Monash University Human Research Ethics Committee 28076); and New Zealand (Health and Disability Ethics Committee 10396). Data may be shared upon reasonable request by contacting the corresponding author.

### Participant eligibility

Inclusion criteria were age ≥35 and ≤75 years; ≥2 ‘poor’ LS7® risk factors self-reported in online screening questionnaire;^20^ no cognitive impairment (Montreal Cognitive Assessment [MoCA] score ³26);^21^ owns a smartphone; and able to speak and understand English. Exclusion criteria were history of stroke, myocardial infarction or terminal illness; participating in another RCT; and family/household members of existing participants.

### Participant recruitment, screening and baseline assessment

Participants were recruited using social media advertising and through primary care organisations (New Zealand only). Interested participants completed an online screening tool to confirm eligibility, except cognitive impairment. If eligible, participants received a link to complete full baseline assessment questionnaires and to make an appointment for a face-to-face assessment of clinical risk factors and cognitive impairment with the MoCA.

Baseline questionnaires elicited details about demographics, stroke awareness, quality of life, health care use, psychological wellbeing, and lifestyle items for the LS7® including diet,^22^ physical activity,^23^ and smoking. The face-to-face physical assessment consisted of height, weight, blood pressure and a point-of-care test (Cardiochek®) for non-fasting cholesterol and glucose.^24^ See supplementary Table 2 (p26) for details.

Participants received a $25 gift card after completing the baseline and 6-month face to face assessments, receiving $50 in total.

### Randomisation

Following the baseline questionnaire and physical assessment, participants were randomised to intervention or usual care group by the project manager via REDCap^25, 26^ using minimisation by site (Auckland, Hamilton, Melbourne, Adelaide, or Hobart).

### Intervention group

The intervention group were sent an e-mail via REDCap providing information on how to download and use the Stroke Riskometer™ App including videos with step-by-step instructions (see Supplement, p13). Participants were e-mailed the necessary health data to complete their information in the App. This contained their individual results obtained from baseline measurements and questionnaires, including each of the LS7® items. One week following randomisation, participants in the intervention group received a three-item online survey via REDCap to ascertain if they had: (1) downloaded; (2) used or (3) required additional assistance required for the App. If people reported needing additional assistance they were contacted by phone by staff not involved in outcome assessments. Only technical support was provided, such as how to download or how to input data into the App, with no information related to their risk factors or results. Reminders about using the App were sent by e-mail at 1 and 3 months after randomisation.

The fidelity of the intervention, that is the use of the App, was intended to be assessed by accessing participants’ user data. The App was designed to only store data on the individual’s device. Participants could choose to share their data with the research team, but this required providing additional consent in a separate section of the App during the set up process. The data available from the App included date of download, individual risk factors and estimated 5- and 10-year risk of stroke. Responses to the 1-week survey and free text responses in the online satisfaction survey completed at 6 months were also used to capture downloads or use of the App.

### Usual care group

The usual care group received an e-mail summary of their risk factors related to the LS7® from their baseline measurements and questionnaire responses with links to online evidence-based stroke prevention information. They were not informed about the App.

### Primary outcome assessment

Follow-up assessments were conducted at 3 months (online), 6 months (face to face) and 12 months (online). The primary outcome was assessed at 6 months using an online questionnaire for LS7 lifestyle items (diet, physical activity, smoking) and a face-to-face assessment including height, weight, blood pressure and point-of-care non-fasting cholesterol and glucose tests by trained research assistants blinded to randomisation group. Before the assessment, participants in the intervention group were reminded not to discuss the App with the research assistant to avoid unblinding. When unblinding did occur, it was recorded as a protocol violation in the REDCap database. There were few harms likely to occur because of this type of app, that is not deemed a medical device per the Therapeutic Goods Administration of Australia. We collected self-reported cardiovascular events by asking about hospitalisations since last contact, which were recorded on the database.

Loss to follow-up of participants in both groups was mostly managed through digital reminders via REDCap. This included e-mails and text messages <3 days before face-to-face appointments. If online questionnaires were not completed, reminders were sent by e-mail at 7 and 14 days. When there was non-response, study staff contacted participants by phone and/or text to encourage participation.

By design there was limited contact between participants and study staff to make the study design closely aligned with ‘real world’ conditions. We also did a process and economic evaluation including collection of additional qualitative data, which will be reported separately.

### Sample size

Assuming a 10% drop-out and 10% drop-in rate, 790 participants (395 intervention, 395 usual care group) provided 80% power (two sided α=0.05) to detect the difference found in the New Zealand pilot in the LS7® of 0.40 (SD 1.61) in the intervention group compared to 0.01 (SD 1.44) in the usual care group at 6 months post-randomisation.^13^ Ongoing monitoring of losses to follow-up demonstrated more drop outs than expected, so recruitment was extended to include 870 people at baseline.

### Primary outcome

The primary outcome was the between-group difference in the change in the LS7® score including blood pressure, non-fasting cholesterol and glucose, BMI, smoking, physical activity, and diet (see Supplementary Table 1 for scoring) from baseline to 6 months post-randomisation.

### Secondary outcomes

A range of secondary outcomes were collected at 3, 6 and 12 months (see Supplementary Table 2, p26). Here we present analyses on the change in individual LS7® items from baseline to 6 months. Other outcomes will be reported separately.

### Analysis

Characteristics of the participants were explored using descriptive statistics.

For the primary outcome analyses we used an analysis of covariance (ANCOVA) to compare the difference in change in total LS7® from baseline to 6 months post-randomisation between the intervention and usual care group. Pre-specified sub-group analyses included examining effect modification by age, sex, country, baseline LS7® and socioeconomic status measured by area-level index of socioeconomic disadvantage. The secondary outcomes were analysed using linear mixed models with adjusted analyses including age, sex, study site and baseline LS7. Sub-group analyses for individual LS7® items were only performed if the main effect of the intervention on the individual LS7® item was p<0.05.

All analyses were conducted using the principle of intention to treat (ITT) by a statistician blinded to randomisation group. Participants who were randomised and who contributed at least one measure of an outcome were included in the intention to treat analysis according to the group to which they were randomised. Multiple imputation using multiple chained equations with 20 imputations imputed missing outcome data using the MICE package in R.^27^ Baseline covariates of treatment group, age, sex, socio-economic status, and country, outcome measures of total LS7® and individual LS7® items (physical activity, diet, smoking, cholesterol, glucose, blood pressure and BMI) and the interaction terms required for subgroup analyses were included in the imputation model.

Sensitivity analyses included a per-protocol analysis. Major protocol violations determined by the Steering Committee included participants who were (1) determined to have not downloaded or used the App based on responses to the 1 week and 6 month surveys; (2) assessed by a research assistant that was unblinded to the treatment allocation; (3) in the usual care group and potentially unblinded due to an administrative error where some participants received an e-mail that referred to the App; and (4) withdrew, lost to follow-up or had incomplete outcome data at 6 months.

Analyses were conducted in R version 4.4.0. All inferences were based on a 5% significance level and two-sided alternatives.

## Results

In total, n=862 participants were recruited from August 2021 to January 2024 with n=433 randomised to the usual care group and n=429 randomised to the intervention group, with per protocol analyses including n=320 in the usual care group and n=276 in the intervention group (Figure 1). The primary outcome assessments at 6 months post-randomisation were completed by the end of July 2024. There were 13 self-reported CVD events (n=7 usual care; n=6 intervention).

**Figure 1.**
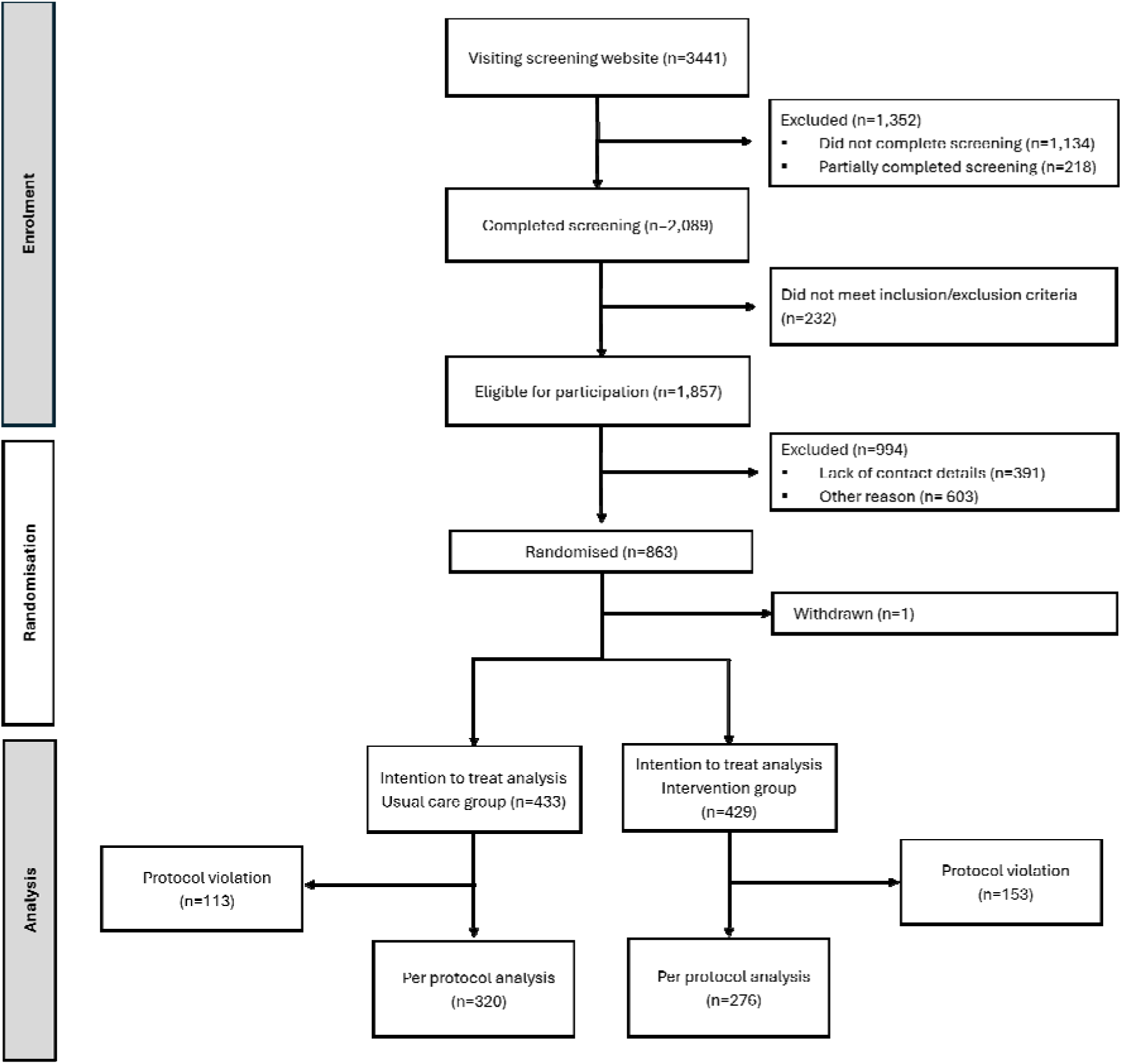
CONSORT flow diagram for the PERKS-International RCT. xxxxxx

The demographic characteristics of the participants of the two groups were balanced although the usual care group had 7% more people in the ‘high’ LS7® group at baseline than the intervention group (Table 1). The individual LS7® items (Supplementary Table 3, p27) were also similar between groups although the intervention group had less people in the ‘ideal’ category for cholesterol (45%) than the usual care group (54%, p=0.02). Less than 10% of people were lost to follow-up and a further 10% were missing one or more LS7® item at 6 months (Supplementary Table 4, p28).

**Table 1.** Demographic characteristics by intervention group.

| Variable | Level | Usual care group<br>n (%) | Intervention group<br>n (%) | Total<br>n (%) |
| --- | --- | --- | --- | --- |
| Total |  | 433 | 429 | 862 |
| Sex | Male | 165 (38%) | 154 (36%) | 319 (37%) |
|  | Female | 268 (62%) | 275 (64%) | 543 (63%) |
| Age category (years) | 35-45 | 72 (17%) | 61 (14%) | 133 (15%) |
|  | 46-55 | 90 (21%) | 115 (27%) | 205 (24%) |
|  | 56-65 | 125 (29%) | 122 (28%) | 247 (29%) |
|  | 66-75 | 146 (34%) | 131 (31%) | 277 (32%) |
| Site | Hobart | 72 (17%) | 74(17%) | 146 (17%) |
|  | Melbourne | 73 (17%) | 72(17%) | 145 (17%) |
|  | Adelaide | 67 (15%) | 132(15%) | 133 (15%) |
|  | Auckland | 189 (44%) | 184 (43%) | 373 (43%) |
|  | Hamilton | 32 (7%) | 33 (8%) | 65 (8%) |
| Ethnicity | European | 314 (73%) | 218 (74%) | 632 (73%) |
|  | Indigenous | 35 (8%) | 40 (9%) | 75 (9%) |
|  | Other* | 84 (19%) | 71 (17%) | 155 (18%) |
| Baseline LS7® category | Low | 90 (21%) | 110 (26%) | 200 (23%) |
|  | Intermediate | 149 (34%) | 155 (36%) | 304 (35%) |
|  | High | 194 (45%) | 164 (38%) | 358 (42%) |
| Baseline total LS7® (mean, SD) |  | 8.13 (1.87) | 7.83 (1.99) | 7.98 (1.93) |
| Area-level SES | 1 (lowest) | 49 (11%) | 78 (18%) | 127 (15%) |
|  | 2 | 67 (15%) | 55 (13%) | 122 (14%) |
|  | 3 | 87 (20%) | 93 (22%) | 180 (21%) |
|  | 4 | 123 (28%) | 109 (25%) | 232 (27%) |
|  | 5 (highest) | 107 (25%) | 94 (22%) | 201 (23%) |
| Self-reported App download in first week |  |  | 277 (64%) |  |
| Self-reported App use in first week |  |  | 264 (62%) |  |
| Downloaded App# |  |  | 339 (79%) |  |

\* includes Asian, South Asian, Islander, African ethnicities.

At 1 week after randomisation, 64% self-reported having downloaded the App and 62% self-reported having used the App including entering their risk factor data. Using data from baseline and follow-up surveys plus user data from the App, we identified 20% of people in the intervention group that we believe did not download or use the App (Supplementary Table 4, p28). Only 32% of people in the intervention group provided additional consent within the App enabling access to their data related to App use. Of these people with App use data, 77% downloaded the App within 14 days of randomisation and 94% downloaded the App within 90 days of randomisation.

The results of the primary outcome analyses with ITT principles and a sensitivity analysis using the per protocol subset, both with multiple imputation of missing data, are shown in Table 2. In the ITT analysis, the mean difference in change from baseline LS7® to follow-up between the intervention and usual care groups was 0.03 (95% CI -0.19, 0.25, p-value 0.79) in the unadjusted model noting within-group improvements in intervention (mean difference 0.18 95% CI 0.02, 0.33) and usual care (mean difference 0.15 95% CI 0.00, 0.29) groups were of similar magnitude. With adjustment for sex, age, country, and baseline LS7®, the results were similar with a mean difference in change from the baseline LS7® to follow-between the intervention and usual care groups of 0.06 (95% CI -0.16, 0.28, p=0.60). The per protocol analysis had a greater, albeit non-significant, effect of the intervention on the change in LS7® over time (unadjusted mean difference in change 0.20 95% CI -0.04, 0.43, p= 0.11; adjusted mean difference in change 0.23 95% CI -0.01, 0.47, p=0.06). Sub-group analyses showed no statistically significant effect of the intervention on the change in LS7® within subgroups defined by age category, sex, country, area-level SES or baseline LS7® (Supplementary Table 5, p29). In addition, tests of interactions in the primary outcome model showed no significant modification of the treatment effect by any of the above factors (all interactions p>0.05).

**Table 2.**
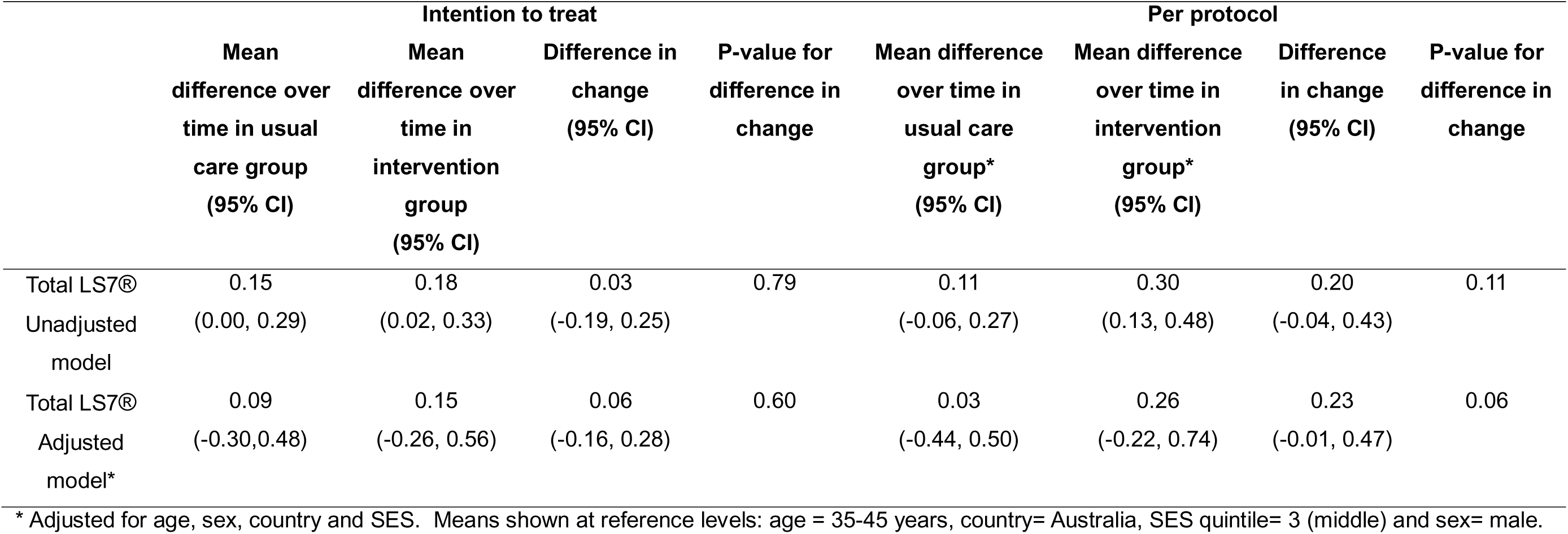
Primary outcome analysis of the differences in the LS7® score from baseline to 6 months post-randomisation in the intervention group compared to the usual care group estimated using ANCOVA using intention to treat and per-protocol samples

The results of the analysis of individual LS7® items are shown in Table 3. Overall, there were no significant differences in the changes in BMI, dietary quality score, blood glucose, cholesterol, systolic blood pressure or current smoking status between the intervention and usual care groups from baseline to 6 months. There was a somewhat greater change in MET-mins/week of physical activity from baseline to 6 months in the intervention than usual care group (mean difference in change in MET-mins/week 313.42 (95% CI -2.80, 629.65, p=0.052)). The magnitude of effect for individual items were consistent in the per protocol analyses. This includes for physical activity (mean difference in change in MET-mins/week 394.24 95% CI 27.80, 760.67, p=0.04; Supplementary Table 6, p30).

**Table 3.** Mean within-person changes in individual LS7® items1 and differences in the mean differences between usual care and interventions groups from baseline to 6-nths

| Item<sup>1</sup> | Adjusted<br>baseline<br>mean*<br>(95% CI) | Mean change over<br>time in usual<br>care group*<br>(95% CI) | Mean change over<br>time in intervention<br>group*<br>(95% CI) | Difference in mean<br>changes between<br>treatment groups*<br>(95% CI) | P-value<br>for<br>difference<br>in change |
| --- | --- | --- | --- | --- | --- |
| BMI (kg/m<sup>2</sup>) | 30.23 (28.55, 31.90) | -0.10 (-0.26, 0.06) | -0.04 (-0.20, 0.12) | 0.06 (-0.17, 0.28) | 0.61 |
| Physical activity (MET-mins/week) | 2075.44 (1524.10,<br>2626.78) | 123.05 (-110.04,<br>356.15) | 436.48 (197.50,<br>675.45) | 313.42 (-2.80, 629.65) | 0.05 |
| Dietary quality score | 9.98 (9.64, 10.33) | 0.19 (0.05, 0.32) | 0.20 (0.06, 0.34) | 0.01 (-0.18, 0.19) | 0.92 |
| Blood glucose (mmol/L) | 5.45 (5.00, 5.89) | 0.05 (-0.15, 0.26) | 0.07 (-0.14, 0.28) | 0.02 (-0.25, 0.29) | 0.89 |
| Systolic blood pressure (mmHg) | 127.84 (124.14, 131.53) | -3.18 (-4.49, -1.86) | -1.93 (-3.27, -0.58) | 1.25 (-0.55, 3.05) | 0.17 |
| Total cholesterol (mmol/L) | 4.44 (4.37, 4.52) | 0.01 (-0.09, 0.11) | 0.05 (-0.05, 0.15) | 0.04 (-0.09, 0.18) | 0.52 |
| Proportion of current smokers | 0.14 (0.10, 0.18) | -0.01 (-0.017, 0.005) | -0.01 (-0.018, 0.004) | -0.0001 (-0.014, 0.014) | 0.99 |

<sup>1</sup> Using only the continuous component of each item without the qualifying factors that are included in the total LS7® calculation. For example, systolic blood pressure in this item analysis is not adjusted according to use of blood pressure medication.
\* Adjusted for age, sex, country and SES. Means shown at reference levels: age category = 35-45 years, country= Australia, SES quintile= 3 (middle) and sex= male.
BMI, body mass index; MET. Metabolic equivalent of Task

## Discussion

This RCT was conducted to examine the efficacy of the Stroke Riskometer™ mHealth application to change the LS7® cardiovascular risk factor score from baseline to 6 months compared to usual care. We were unable to detect any difference in the change in the overall LS7® score between the intervention and usual care groups over time with results consistent among sub-groups of age, sex, area level SES, baseline LS7® and country.

There was some indication of a greater effect of the App in per protocol than ITT analyses, but these were still very small and not statistically significant, consistent with the ITT. There was generally no association between randomisation to use of the App and individual LS7® items; although, there was some evidence of a greater improvement in MET-mins of physical activity between baseline and 6 months for the intervention group compared to the usual care group. The limited associations could be related to the Stroke Riskometer™ mHealth app or due to aspects of the study design and delivery, which the process evaluation, to be reported separately, will explore further Our study is one of the few fully-powered, mHealth RCTs with objectively measured risk factors focused on a primary prevention population.^28^ Although it is difficult to directly compare our findings to others given the heterogeneity of mHealth apps, authors of other RCTs have similarly demonstrated no association between use of an mHealth intervention and changes in objectively measured cardiovascular risk factors,^29, 30^ particularly in a primary prevention setting.^28^ In contrast, in a recent study in India in people with prediabetes or obesity a more comprehensive, structured 12 week app-based intervention including telephone health coaching was reported to improve weight, waist circumference and blood pressure but not lipids after 3 months.^31^ There is somewhat greater evidence that mHealth apps may be effective in people with pre-existing CVD.^28^ For example, in a recent review 8 out of 14 mHealth RCTs were found to have positive changes in blood pressure; 7 out of 12 had improvements in weight and 4 out of 11 had improvements in lipids in intervention compared to control groups.^28^ The somewhat, although not unequivocally, greater effects of mHealth interventions in people with a history of CVD may be attributable to differences in motivation and engagement. Having an acute cardiovascular event is proposed in some behaviour change models as a ‘teachable moment’ that can lead to positive changes in health behaviour,^32, 33^ which is not present in a general population as included in our trial.

There was a marginally greater increase in MET-minutes of physical activity per week, as measured using a validated questionnaire, in the intervention than usual care groups. Given the lack of effect on the LS7® score overall and other LS7® items, as well as the wide confidence interval, these findings should be interpreted with caution. The App does provide specific feedback and advice to people who report low levels of physical activity, including a push notification about doing physical activity when people identified increasing physical activity as a goal. Of note is that the App has the same features for smoking, diet, weight, stress and alcohol consumption that did not improve in the intervention group. In support of the finding, there is other evidence that mHealth apps can increase self-reported and objectively measured physical activity,^11^ although the intensity of those interventions that mostly focused on physical activity specifically appears greater than ours.

The lack of an effect of the Stroke Riskometer App on the combined LS7® outcome could be, in part, due to the design and conduct of our trial. We took a real-world approach, with minimal interaction between participants and study staff. Reminders mostly came via e-mail and text messages with phone contact used as the final option. While these processes meant our study reflects a person downloading and using the App in the ‘real world’ these processes may have limited the fidelity of the intervention and overall participant engagement. However, additional face to face or telephone support about the App could have become a co-intervention, potentially masking the effect of the App alone. In support of our study design and implementation, over two thirds of people self-reported downloading the app within 1 week, this being before we did any targeted engagement with participants and less than 10% of people were lost to follow-up. However, 20% of people in the intervention group were determined to have not downloaded or used the App at 6 months.

The numerically larger, but still not statistically significant, intervention effect for the overall LS7® score and some individual items in the per protocol compared to ITT analysis provides some evidence that the lack of fidelity of the intervention delivery has played a role in our null findings.

The findings may also be explained by aspects of the intervention itself. While the App incorporates some evidence-based behaviour change tools it does not include more recent advances in mobile technology that may increase engagement and effectiveness. Advances such as gamification, interactive components such as chat bots using generative artificial intelligence and integration of data from wearables, e.g. blood pressure or physical activity, could improve the Stroke Riskometer’s™ utility.^34^ Since completion of the trial, the App has been updated to enhance the user experience, including providing more specific feedback on each risk factor. An embedded quantitative and qualitative process evaluation is pending that will explore the experience of trial participants. Information from the evaluation will be used to update the app to potentially improve its effectiveness on risk factor change.

The major limitations are that 20% of study participants in the intervention group did not download the app during the study and 38% did not download it within one week after randomisation. We also lacked detailed information on the fidelity of the intervention because less than a third of participants provided the additional consent required to access App user information. While this is a strength of the App because it means that users’ data are secure on their device, it is a disadvantage for our RCT because this consent process required several additional steps inside the App. The public availability of the App may be considered a limitation. To avoid contamination of the usual care group, we could not promote that the trial was of a stroke prevention App. While we advertised that the study would involve receiving information on a mobile phone, we likely recruited people who were not interested in an App-based intervention. This could be considered a strength of our study, as we have replicated a real-world setting where the general population may be offered an app to manage their health. We used the American Heart Association’s LS7® to assess risk factors, which is now superseded by the Life’s Essential 8 including sleep and, most recently, Life’s Crucial 9.^35^ We did not have sleep data so could not explore these scores as an outcome. A further consideration is the improvement in the LS7® from baseline to follow-up for the usual care group, which dilutes the difference in change in LS7® between groups.

The usual care group received an e-mail with results of the comprehensive risk factor assessment and links to evidence-based information on managing these risk factors once after randomisation. This appears to have resulted in a small (e.g. 0.15 units from a range of 0 to 14 units for the total score) but nonetheless positive change in risk factors. The strengths of the study include the diversity of participants including people from different ethnic groups in Australia and New Zealand – both being multicultural countries – and a good balance of males, females and different age groups. The objective measures of blood pressure, glucose, lipids and weight at baseline and follow-up are strengths, with mHealth studies often only focusing on self-reported risk factors. Other strengths were meeting our recruitment target, low losses to follow-up and limited protocol violations meaning our analyses are fully powered.

In conclusion, in a real-world setting, providing information on stroke risk factors and access to a personalised mHealth app the Stroke Riskometer™ about stroke and its risk factors does not result in overall benefit to stroke risk factor profile after 6 months compared to a usual care group. To better leverage mHealth applications to reduce stroke risk, Apps may have to include more features to promote engagement and be embedded in models incorporating face to face or telehealth support in health care settings.

## Data Availability

Data may be shared upon reasonable request by contacting the corresponding author.

## Acknowledgements and funding

This study was funded by the National Health and Medical Research Council in Australia through Synergies TO Prevent Stroke (STOPstroke) Synergy Grant (GNT1182071). SLG is funded by a National Heart Foundation of Australia Future Leader Fellowship (108524). We would like to thank the members of the STOPstroke Scientific Advisory Committee for their oversight of the RCT including Dr Eleanor Horton (chair), Professor Graeme Hankey, Professor Chris Sobey, Professor Julie Redfern, Dr Bo Reminyi, and Ms Miriam Lum On.

## References

1. Feigin VL, Krishnamurthi R, Nair B, Rautalin I, Parag V, Anderson CS, et al. Trends in stroke incidence, death, and disability outcomes in a multi-ethnic population: Auckland regional community stroke studies (1981-2022). Lancet Reg Health West Pac 2025; 56: 101508.

2. O’Donnell MJ, Chin SL, Rangarajan S, Xavier D, Liu L, Zhang H, et al. Global and regional effects of potentially modifiable risk factors associated with acute stroke in 32 countries (INTERSTROKE): a case-control study. Lancet 2016; 388: 761–775.

3. Australian Bureau of Statistics. National Health Survey, https://www.abs.gov.au/statistics/health/health-conditions-and-risks/national-health-survey/2022 (2023, accessed 7th May 2025).

4. Ministry of Health New Zealand. New Zealand Health Survey 2023/24. 2024. Wellington, New Zealand.

5. National Centre for Health Statistics. Hypertension Prevalence, Awareness, Treatment and Control among adults aged 18 years and older: United States August 2021-August 2023. 2024. USA.

6. National Centre for Health Statistics. Obesity and Severe Obesity Prevalence in Adults: United States, August 2021–August 2023. 2024. USA.

7. National Health Service. Health Survey for England 2022, Part 2. 2024. UK.

8. Stroke Association. Shaping stroke research to rebuild lives: The Stroke Priority Setting Partnership results for investment. 2021.

9. Shanahan M and Bahia K. GSMA The State of Mobile Internet Connectivity 2024. 2024.

10. Abe M, Hirata T, Morito N, Kawashima M, Yoshida S, Takami Y, et al. Smartphone application-based intervention to lower blood pressure: a systematic review and meta-analysis. Hypertension Research 2025; 48: 492–505.

11. Bushey E, Wu Y, Wright A and Pescatello L. The Influence of Physical Activity and Diet Mobile Apps on Cardiovascular Disease Risk Factors: Meta-Review. J Med Internet Res 2024; 26: e51321.

12. Shi B, Li G, Wu S, Ge H, Zhang X, Chen S, et al. Assessing the Effectiveness of eHealth Interventions to Manage Multiple Lifestyle Risk Behaviors Among Older Adults: Systematic Review and Meta-Analysis. J Med Internet Res 2024; 26: e58174.

13. Krishnamurthi R, Hale L, Barker-Collo S, Theadom A, Bhattacharjee R, George A, et al. Mobile technology for primary stroke prevention. Stroke 2019; 50: 1–3.

14. Michie S, Richardson M, Johnston M, Abraham C, Francis J, Hardeman W, et al. The Behavior Change Technique Taxonomy (v1) of 93 Hierarchically Clustered Techniques: Building an International Consensus for the Reporting of Behavior Change Interventions. Annals of Behavioral Medicine 2013; 46: 81–95.

15. Lloyd-Jones DM, Hong Y, Labarthe D, Mozaffarian D, Appel LJ, Van Horn L, et al. Defining and setting national goals for cardiovascular health promotion and disease reduction: the American Heart Association’s strategic Impact Goal through 2020 and beyond. Circulation 2010; 121: 586–613.

16. Gall SL, Feigin V, Thrift AG, Kleinig TJ, Cadilhac DA, Bennett DA, et al. Personalized knowledge to reduce the risk of stroke (PERKS-International): Protocol for a randomized controlled trial. Int J Stroke 2022: 17474930221113430.

17. Hopewell S, Chan AW, Collins GS, Hróbjartsson A, Moher D, Schulz KF, et al. CONSORT 2025 statement: updated guideline for reporting randomised trials. Bmj 2025; 389: e081123.

18. Hoffmann TC, Glasziou PP, Boutron I, Milne R, Perera R, Moher D, et al. Better reporting of interventions: template for intervention description and replication (TIDieR) checklist and guide. BMJ : British Medical Journal 2014; 348: g1687.

19. Agarwal S, LeFevre AE, Lee J, L’engle K, Mehl G, Sinha C, et al. Guidelines for reporting of health interventions using mobile phones: mobile health (mHealth) evidence reporting and assessment (mERA) checklist. bmj 2016; 352.

20. Lloyd-Jones Donald M, Hong Y, Labarthe D, Mozaffarian D, Appel Lawrence J, Van Horn L, et al. Defining and Setting National Goals for Cardiovascular Health Promotion and Disease Reduction. Circulation 2010; 121: 586–613.

21. Nasreddine ZS, Phillips NA, Bédirian V, Charbonneau S, Whitehead V, Collin I, et al. The Montreal Cognitive Assessment, MoCA: A brief screening tool for mild cognitive impairment. Journal of the American Geriatrics Society 2005; 53: 695–699.

22. Cleghorn CL, Harrison RA, Ransley JK, Wilkinson S, Thomas J and Cade JE. Can a dietary quality score derived from a short-form FFQ assess dietary quality in UK adult population surveys? Public Health Nutrition 2016; 19: 2915–2923.

23. Craig CL, Marshall AL, Sjostrom M, Bauman AE, Booth ML, Ainsworth BE, et al. International physical activity questionnaire: 12-country reliability and validity. Med Sci Sports Exerc 2003; 35: 1381–1395.

24. Robert Lourdes TG, Chong ZL, Saminathan TA, Abd Hamid HA, Mat Rifin H, Wan KS, et al. Diagnostic accuracy of Cardiochek(®) PA point-of-care testing (POCT) analyser with a 3-in-1 lipid panel for epidemiological surveys. Lipids Health Dis 2024; 23: 297.

25. Harris PA, Taylor R, Minor BL, Elliott V, Fernandez M, O’Neal L, et al. The REDCap consortium: Building an international community of software platform partners. Journal of Biomedical Informatics 2019; 95: 103208.

26. Harris PA, Taylor R, Thielke R, Payne J, Gonzalez N and Conde JG. Research electronic data capture (REDCap)—A metadata-driven methodology and workflow process for providing translational research informatics support. Journal of Biomedical Informatics 2009; 42: 377–381.

27. White IR, Royston P and Wood AM. Multiple imputation using chained equations: Issues and guidance for practice. Stat Med 2011; 30: 377–399.

28. Qi Y, Mohamad E, Azlan AA, Zhang C, Ma Y and Wu A. Digital Health Solutions for Cardiovascular Disease Prevention: Systematic Review. J Med Internet Res 2025; 27: e64981.

29. Morawski K, Ghazinouri R, Krumme A, Lauffenburger JC, Lu Z, Durfee E, et al. Association of a Smartphone Application With Medication Adherence and Blood Pressure Control: The MedISAFE-BP Randomized Clinical Trial. JAMA internal medicine 2018; 178: 802–809.

30. Yousuf H, Reintjens R, Slipszenko E, Blok S, Somsen GA, Tulevski II, et al. Effectiveness of web-based personalised e!ZICoaching lifestyle interventions. Netherlands Heart Journal 2019; 27: 24–29.

31. Muralidharan S, Ranjani H, Anjana RM, Gupta Y, Ambekar S, Koppikar V, et al. Change in cardiometabolic risk factors among Asian Indian adults recruited in a mHealth-based diabetes prevention trial. DIGITAL HEALTH 2021; 7: 20552076211039032.

32. Brust M, Gebhardt WA, Ter Hoeve N, Numans ME and Kiefte-de Jong JC. Exploring timing and delivery of lifestyle advice following an acute cardiac event hospitalization: The cardiac patient’s perspective. Patient Educ Couns 2024; 124: 108279.

33. McBride CM, Emmons KM and Lipkus IM. Understanding the potential of teachable moments: The case of smoking cessation. Health Education Research 2003; 18: 156–170.

34. Zhang R and Wang H. Insights into the Technological Evolution and Research Trends of Mobile Health: Bibliometric Analysis. Healthcare (Basel) 2025; 13.

35. Gaffey AE, Rollman BL and Burg MM. Strengthening the Pillars of Cardiovascular Health: Psychological Health is a Crucial Component. Circulation 2024; 149: 641–643.

