## Supplementary materials for "PERsonalised Knowledge to reduce the risk of Stroke (PERKS-International): a randomised controlled trial testing the efficacy of an mHealth application to reduce risk factors for the primary prevention of stroke"

Supplementary materials contents

### Supplementary material 1 - CONSORT 2025 expanded checklist of information to include when reporting a randomised trial*

| Section/Topic | | Item No | | Checklist item | | Reported on page No |
| --- | --- | --- | --- | --- | --- | --- |
| Title and structured abstract | | | | | | |
|  | | 1a | | Identification as a randomised trial in the title | | 1 |
|  |  | 1b | | Structured summary of trial design, methods, results, and conclusions (for specific guidance see CONSORT for abstracts) | | 3 |
| Open Science |  | |  | |  | |
| Trial registration | 2 | | Name of trial registry, identifying number (with URL) and date of registration | | 6 | |
| Protocol and statistical analysis plan | 3 | | Where the trial protocol and statistical analysis plan can be accessed | | Reference 12 | |
| Data sharing | 4 | | Where and how the individual deidentified participant data (including data dictionary), statistical code and any other materials can be accessed | | 7 | |
| Funding and conflicts of interest | 5a | | Sources of funding and other support (e.g., supply of drugs), and role of funders in the design, conduct, analysis and reporting of the trial | | 27 | |
|  | 5b | | Financial and other conflicts of interest of the manuscript authors | | 27 | |
| Introduction | | | | | | |
| Background and objectives | | 6 | | Scientific background and explanation of rationale | | 5-6 |
|  |  | 7 | | Specific objectives related to benefits and harms | | 6 |
| Methods | | | | | | |
| Patient and public involvement | 8 | | Details of patient or public involvement in the design, conduct and reporting of the trial | | 6-7 | |
| Trial design | | 9 | | Description of trial design (such as parallel, factorial) including allocation ratio | | 6 |
| Changes to trial protocol | | 10 | | Important changes to methods after trial commencement (such as eligibility criteria), with reasons | | N/A |
| Trial setting | | 11 | | Settings (e.g., community, hospital) and locations (e.g., countries, sites) where the trial was conducted | | 6 |
| Eligibility criteria | | 12a | | Eligibility criteria for participants | | 7 |
|  |  | 12b | | If applicable, eligibility criteria for sites and for individuals delivering the interventions (e.g., surgeons, physiotherapists) | | N/A |
| Intervention and comparator | | 13 | | Intervention and comparator with sufficient details to allow replication. If relevant, where additional materials describing the intervention and comparator (e.g., intervention manual) can be accessed | | 5, 6, 8, 9 |
| Outcomes | | 14 | | Pre-specified primary and secondary outcomes | | 9-10 |
| Harms | | 15 | | How harms were defined and assessed (e.g., systematically, nonsystematically) | | 9 |
| Sample size | | 16a | | How sample size was determined | | 10 |
|  |  | 16b | | When applicable, explanation of any interim analyses and stopping guidelines | | N/A |
| Randomisation: | |  | |  | |  |
| Sequence generation | | 17a | | Who generated the random allocation sequence and method used | | 8 |
|  |  | 17b | | Type of randomisation; details of any restriction (such as blocking and block size) | | 8 |
| Allocation concealment mechanism | | 18 | | Mechanism used to implement the random allocation sequence (such as sequentially numbered containers), describing any steps taken to conceal the sequence until interventions were assigned | | 8 |
| Implementation | | 19 | | Whether the personnel who enrolled and those who assigned participants to the interventions had access to the random allocation sequence | | 8 |
| Blinding | | 20a | | Who was blinded after assignment to interventions (e.g., participants, care providers, outcome assessors, data analysts) | | 8 and 9 |
|  |  | 20b | | If blinded, how blinding was achieved and description of the similarity of interventions | | 8 and 9 |
| Statistical methods | | 21a | | Statistical methods used to compare groups for primary and secondary outcomes | | 11 |
|  |  | 21b | | Definition of who is included in each analysis (e.g., all randomised participants), and in which group | | 11 and 12 |
|  | | 21c | | How missing data were handled in the analysis | | 11 |
|  | | 21d | | Methods for any additional analyses (e.g., subgroup and sensitivity analyses), distinguishing prespecified from post-hoc | | 11-12 |
| Results | | | | | | |
| Participant flow, including flow diagram | | 22a | | For each group, the numbers of participants who were randomly assigned, received intended treatment, and were analysed for the primary outcome | | 11 and Figure 1 |
|  |  | 22b | | For each group, losses and exclusions after randomisation, together with reasons | | 13 and supplementary table 4 |
| Recruitment | | 23a | | Dates defining the periods of recruitment and follow-up outcomes of benefits and harms | | 12 |
|  |  | 22b | | Why the trial ended or was stopped | | N/A |
| Intervention and comparator delivery | | 24a | | Intervention and comparator as they were actually administered (e.g., where appropriate, who delivered the intervention/comparator, whether participants adhered, whether they were delivered as intended [fidelity]) | | 14 |
|  | | 24b | | Concomitant care received during the trial for each group | | N/A |
| Baseline data | | 25 | | A table showing baseline demographic and clinical characteristics for each group | | 15, Table 1 |
| Numbers analysed | | 26 | | For each primary and secondary outcome, by group:● the number of participants included in the analysis ● the number of participants with available data at the outcome time point ● result for each group, and the estimated effect size and its precision (such as 95% confidence interval) ● for binary outcomes, presentation of both absolute and relative effect size | | Figure 1, Table 3  16 and Table 2  18 and Table 3 |
| Harms | | 27 | | All important harms or unintended effects in each group (for specific guidance see CONSORT for harms) | | 12 |
| Ancillary analyses | | 28 | | Results of any other analyses performed, including subgroup analyses and adjusted analyses, distinguishing pre-specified from exploratory | | 16 and 18, Table 2 and Supplementary Tables 5 and 6 |
| Discussion | | | | | | |
| Interpretation | | 22 | | Interpretation consistent with results, balancing benefits and harms, and considering other relevant evidence | | 20-22 |
| Limitations | | 20 | | Trial limitations, addressing sources of potential bias, imprecision, and, if relevant, multiplicity of analyses | | 22-23 |

Supplementary material 2 - The TIDieR (Template for Intervention Description and Replication) Checklist***:**

Information to include when describing an intervention and the location of the information

| **Item number** | **Item** | **Where located **** | |
| --- | --- | --- | --- |
|  |  | Primary paper  (page or appendix  number) | Other ^†^ (details) |
|  | **BRIEF NAME** |  |  |
| **1.** | Provide the name or a phrase that describes the intervention. | _____5_______ | ______________ |
|  | **WHY** |  |  |
| **2.** | Describe any rationale, theory, or goal of the elements essential to the intervention. | 5, 6 and 8______ | Protocol paper: https://doi.org/10.1177/17474930221113430 |
|  | **WHAT** |  |  |
| **3.** | Materials: Describe any physical or informational materials used in the intervention, including those provided to participants or used in intervention delivery or in training of intervention providers. Provide information on where the materials can be accessed (e.g. online appendix, URL). | 5, 8, Supplement and google play or apple app store for free download of app | Protocol paper: https://doi.org/10.1177/17474930221113430 |
| **4.** | Procedures: Describe each of the procedures, activities, and/or processes used in the intervention, including any enabling or support activities. | 8, supplement for ‘how to’ video scripts | _____________ |
|  | **WHO PROVIDED** |  |  |
| **5.** | For each category of intervention provider (e.g. psychologist, nursing assistant), describe their expertise, background and any specific training given. | __8__________ | _____________ |
|  | **HOW** |  |  |
| **6.** | Describe the modes of delivery (e.g. face-to-face or by some other mechanism, such as internet or telephone) of the intervention and whether it was provided individually or in a group. | ___5 and 8____ | _____________ |
|  | **WHERE** |  |  |
| **7.** | Describe the type(s) of location(s) where the intervention occurred, including any necessary infrastructure or relevant features. | 5 and 8_ | _____________ |
|  | **WHEN and HOW MUCH** |  |  |
| **8.** | Describe the number of times the intervention was delivered and over what period of time including the number of sessions, their schedule, and their duration, intensity or dose. | 8, 14, Table 1_ | _____________ |
|  | **TAILORING** |  |  |
| **9.** | If the intervention was planned to be personalised, titrated or adapted, then describe what, why, when, and how. | __5 and 6___ | _____________ |
|  | **MODIFICATIONS** |  |  |
| **10.^ǂ^** | If the intervention was modified during the course of the study, describe the changes (what, why, when, and how). | __N/A____ | _____________ |
|  | **HOW WELL** |  |  |
| **11.** | Planned: If intervention adherence or fidelity was assessed, describe how and by whom, and if any strategies were used to maintain or improve fidelity, describe them. | _8, 9 | _____________ |
| **12.^ǂ^** | Actual: If intervention adherence or fidelity was assessed, describe the extent to which the intervention was delivered as planned. | 14_ | _____________ |

** **Authors** - use N/A if an item is not applicable for the intervention being described. **Reviewers** – use ‘?’ if information about the element is not reported/not sufficiently reported.

† If the information is not provided in the primary paper, give details of where this information is available. This may include locations such as a published protocol or other published papers (provide citation details) or a website (provide the URL).

ǂ If completing the TIDieR checklist for a protocol, these items are not relevant to the protocol and cannot be described until the study is complete.

* We strongly recommend using this checklist in conjunction with the TIDieR guide (see *BMJ* 2014;348:g1687) which contains an explanation and elaboration for each item.

* The focus of TIDieR is on reporting details of the intervention elements (and where relevant, comparison elements) of a study. Other elements and methodological features of studies are covered by other reporting statements and checklists and have not been duplicated as part of the TIDieR checklist. When a **randomised trial** is being reported, the TIDieR checklist should be used in conjunction with the CONSORT statement (see [www.consort-statement.org](http://www.consort-statement.org)) as an extension of **Item 5 of the CONSORT 2010 Statement.** When a **clinical trial** **protocol** is being reported, the TIDieR checklist should be used in conjunction with the SPIRIT statement as an extension of **Item 11 of the SPIRIT 2013 Statement** (see [www.spirit-statement.org](http://www.spirit-statement.org)). For alternate study designs, TIDieR can be used in conjunction with the appropriate checklist for that study design (see [www.equator-network.org](http://www.equator-network.org)).

### Supplementary material 3 - mHealth evidence reporting and assessment (mREA) guidelines

*Infrastructure – item 1*

The study is based in Australia and New Zealand. In Australia there are three main mobile network providers, these have 99.4%, 98.5% and 96% coverage of the Australian population. In New Zealand, there are also three main mobile network providers, these have coverage based on where people in the country work and recreate with all reporting coverage of 98.5%.

Information reported on page 5 of main text.

*Technology/platform – item 2*

The App was developed by researchers in conjunction with Auckland University of Technology (AUT) Ventures as specified in the disclosures section of the manuscript (page 27). It Is developed for iOS and Android platforms. React Native programming language is used for both platforms. It is available for free in Google Play and the Apple App Store. The algorithm the provides relative and absolute risk of stroke to users was developed and validated by researchers at the Auckland University of Technology.^1^

*Interoperability – item 3*

The App is designed for use by the general community. It does not directly interface with health information systems. Users can download their results and progress, which can be shared with health care providers.

*Intervention delivery – item 4*

Details on the App are provided in introduction (page 5 and 6) and methods (page 8) of the main text. Details on delivery of intervention and control group are given on page 8 and 9 of the main text.

*Intervention content – item 5*

Introduction on page 5, methods on page 8 and supplementary information (page 9 and 10 to 14).

*Usability and content testing – item 6*

Reported in supplement, page 11.

*User feedback – item 7*

Supplementary material (page 11). A process evaluation was conducted as part of the RCT (page 10) but will be reported separately.

*Access of individual participants – item 8*

Supplemental material (page 11).

*Cost assessment – item 9*

Economic evaluation to be undertaken as part of secondary outcomes (page 9).

*Adoption inputs/programme entry – item 10*

Recruitment strategies and information provided to participants randomised to intervention group with app are included on page 7 and 8 of manuscript.

*Limitations for delivery at scale – item 11*

App is free and available in multiple (19) languages facilitating scale up, see Page 6.

*Contextual adaptability – item 12*

App is available in 19 languages, see supplement page 11.

App is updated regularly based on user feedback, which can be submitted through the relevant app stores or through the e-mail address provided in the app in supplemental. See also page 20 and 21 of manuscript.

Process evaluation to identify adaptation to app, see page 9 of manuscript.

*Replicability – item 13*

App is available for free to download to allow others to use and/or test the app in different settings, page 6 of manuscript.

*Data security – item 14*

See supplement page 11.

*Compliance with national guidelines/regulations – item 15*

Page 6 and 7 of manuscript.

*Fidelity of intervention – item 16*

Page 8, page 10, page 14, Table 1.

### Supplementary material 4 - Stroke Riskometer mobile phone application

Technology

The App was developed by researcher in conjunction with AUT Ventures. It Is developed for iOS and Android platforms. React Native programming language is used for both platforms, which is quite common and well known. It is available for free in Google Play and the Apple App Store. The algorithm the provides relative and absolute risk of stroke to users was developed and validated by researchers at the Auckland University of Technology.^1^

Usability and content testing

The App was developed in conjunction with consumers including health professionals and the general public. This process included a series of focus groups and discussions before development and initial launch in 2014.

Users can provide feedback to the developers via the Google Play or Apple App Store and the e-mail function with the app itself. A pilot RCT (reference) also captured user feedback on the App. These feedback mechanisms have resulted in over 200 modification to the App since its launch in 2014.

Contextual adaptability

The App is available in 25 languages. The App is updated regularly based on consumer feedback provided through the app store and direct communication via e-mail with the developers. To date there have been over 200 modifications to the app based on feedback.

Data security

The App is protected by Hypertext Transfer Protocol Secure (https) protocols. Users data is not shared with the App developers unless the user selects to be a part of the optional Reducing the International Burden of Stroke Using Mobile Technology (RIBURST) study. The study is conducted according to the GCP Guidelines and relevant regional regulatory (Ethics) Committee requirements. Participation is completely voluntary. The study is reachable through the Research section of the Stroke Riskometer app. If the user will agree to participate in the RIBURST and, their data will be submitted and will be kept in a secure database located on premises at AUT in New Zealand; a country which the European Union Commission has deemed as having an adequate level of data protection. All communication between the app and the data collection services are encrypted using https protocol. The data will be preserved by technical, physical and administrative measures from loss, unauthorised disclosure or change.

### Supplementary material 5 - Description of instructional videos provided to intervention group participants

Instructional videos were created by a media professional and included animations and videos with voice overs explaining key steps in downloading and using the Stroke Riskometer app. These were provided to intervention group participants in the e-mail after randomisation as links. They were hidden from public view.

*Video about the App – animation or clips with voice over by local people (e.g. Australian and New Zealand)*

- The Stroke Riskometer App has been designed by stroke experts to help you reduce your risk of stroke
- The App is endorsed by the international bodies including the World Federation of Neurologists, The World Stroke Organisation, the European Stroke Organisation and the World Heart Federation
- To download the App you can visit either the App store or Google play depending on the type of phone you have.
- Follow the link in the e-mail we sent you for either the iPhone [show image of iPhone] or android devices [show images of Samsung galaxy or google phone].
- Once you are in the App store on the Stroke Riskometer page click ‘get’. You may need to double click the button on the right of your screen depending on the type of phone you have.
- The phone will now show you it is downloading the app.
- Once it has finished downloading click ‘open’.
- You will now be inside the Stroke Riskometer App.
- The app has 4 sections that we will talk about more in further videos.
- These are ‘assess’, which gets you to enter your risk factors, ‘research’, which will register the app to you for research purposes, ‘manage’ that will empower you with information about your risk and how to change them, and ‘FAST’ which will educate you about the signs and symptoms of stroke.
- The app will send you notification to remind you to use it and of your goals.

*Video about how to enter information into the App*

- The Stroke Riskometer App uses your information to provide you with personalised information about your relative and absolute risk of stroke
- When you open the app for the first time you will see the home screen. You should click ‘Begin test’.
- Follow the instructions on the screen completing your risk factor information. We have provided all of your answers in the e-mail we sent to you from the assessments we did the other day.
- If you want more information about any of the questions, click on the ‘information’ button.
- At the bottom of each screen, your estimated risk of stroke is shown. Click ‘next’ to move between screens.
- If you need to go back, press the back arrow in the top left hand corner.
- When you have entered all of the risk factors on page 7 of 7 press ‘calculate’
- You can select to see your results as a ‘bar’ [show bar option] or as a ‘dial’ [show dial option].
- In the bar option, you can see your current risk level for having a stroke in next 5 or 10 years. The red bar shows your current level of risk based on your risk factors. The green bar shows what your risk of stroke would be if you reduced all of your risk factors to healthy levels.
- If the red bar is bigger than the green bar you can makes changes to your lifestyle and reduce your risk of stroke.
- In the dial option, you can see your current risk level for having a stroke in next 5 or 10 years. The text describes what is meant by your relative risk compared to people who do not have the same risk factors as you. This risk could be reduced by changing your risk factors.
- Click ‘save your results’. When the box pops up press ‘yes’.
- You also need to click ‘participate in research’. This will allow us to follow your use of the app. Confirm that the information you have supplied is correct. Then specify that you are a ‘new user’. Read the information sheet and check the boxes to show you understand.
- Finally you enter your name, e-mail address, password and an alternative e-mail address to register the app to you.

*Video about managing your risk with the App*

- You can revisit your risk factor information at any time.
- If you open the app, you will see the bar at the bottom that has 4 buttons.
- Click ‘manage’ to open the risk factor section of the app.
- Now you can see your risk factors for stroke on this screen. You identified risk factors are on the top section in yellow.
- Click the arrow to get further information.
- You can also navigate to the video section to see other information about this risk factor.
- We suggest that you spend some time reading this information over the next few weeks. The App will also send you reminders to get you to check in with the app regularly.
- In the ‘manage your risk’ home page, press ‘your goals’. In here you should select options for the risk factors you would like to change most.
- For example, we can select ‘stop smoking’, and to get a daily reminder about this.
- If you take medication you can also set reminders in the App so you don’t forget.
- For example, if I take Lipitor one a day. You can type ‘lipitor’ and then set the time you would like to take it and what days you take it.
- To get back to the home section of the ‘manage your risk’ section you can just click ‘manage’

*Video about tracking your progress over time*

- The Riskometer app will allow you to re-enter your risk factors so that you can monitor how your stroke risk has changed.
- To do this, open the app and click assess. You can re-enter all of the risk factors changing these where necessary.
- For example, if you cut down your smoking you could select a different category here.
- Or if you lost weight, you can adjust the weight.
- Change as many risk factors as is relevant. Then click through to ‘calculate’
- You will now see your updated risk in either bar or dial format.
- Make sure you hit ‘save your results!’ and then indicate ‘yes’ for the pop up box about the progress chart.
- Now if we go to the manage section and ‘progress graphs’ we can see our overall stroke risk and individual risk factors.
- Seeing positive changes can help motivate you to continue with your good habits.

*Video about the FAST section of the app*

- The Riskometer app also provided important information about the signs and symptoms of stroke
- Click on ‘FAST’ on the bottom of the screen.
- Here you will see detailed information about how to recognise a stroke and what to do.
- “F” is for Face drooping
- “A” is for Arm weakness
- “S” is for Speech difficulty
- “T” is for Time. It is important to call an ambulance if the person shows any of these symptoms. You should still call an ambulance even if the symptoms go away very quickly.

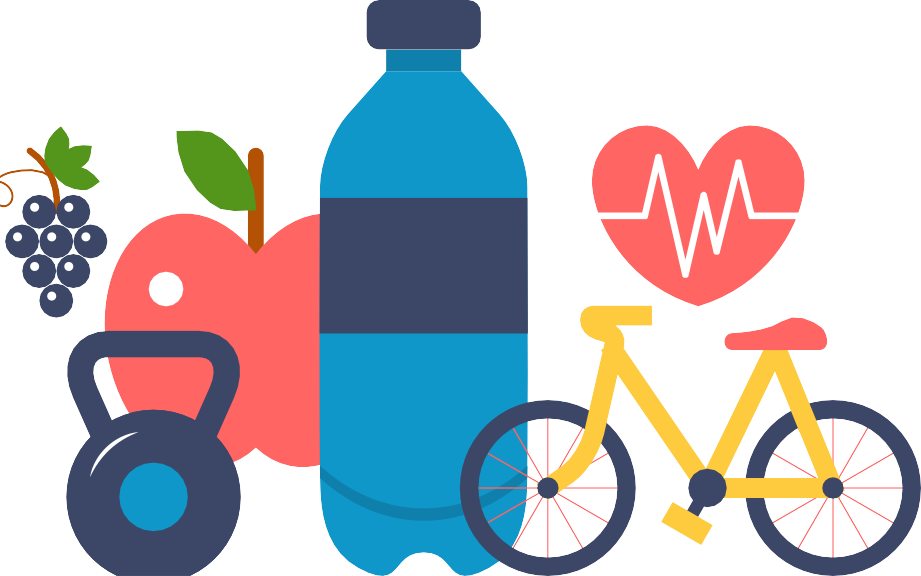

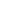

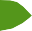

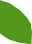

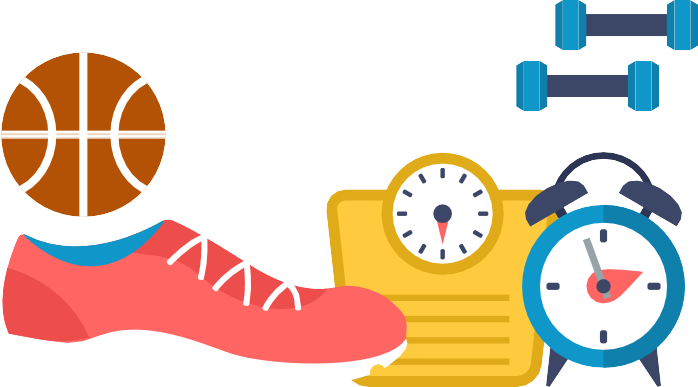

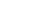

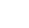

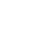

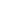

### Supplementary material 6 - PERsonalised Knowledge to reduce the risk of Stroke (PERKS-International Trial) Participant Information Sheet and consent

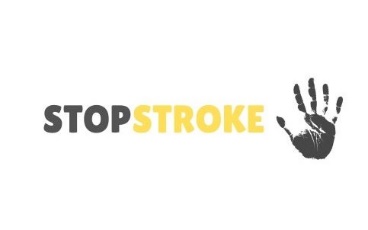

| Research team | Associate Professor Seana Gall*,*  Menzies Institute for Medical Research, University of Tasmania   Professor Timothy Kleinig  Royal Adelaide Hospital   Professor Amanda Thrift  Monash University   Professor Dominique Cadilhac  Monash University  |
| --- | --- |

**Invitation**

You are invited to take part in a clinical trial titled “PERsonalised Knowledge to reduce the risk of Stroke (PERKS-International)”. Your participation in this study will be for a duration of 12 months. This study is coordinated by the University of Tasmania with collaboration from the Auckland University of Technology, Monash University and the Royal Adelaide Hospital (Central Adelaide Local Health Network).

This Participant Information Sheet/Consent Form tells you about the research project. It explains the tests involved. Knowing what is involved will help you decide if you want to take part in the research. Please read this information carefully. Ask questions about anything that you don’t understand or want to know more about. Before deciding whether or not to take part, you might want to talk about it with a relative, friend or your local doctor.

**What is the purpose of this study?**

Stroke is one of the biggest causes of death and disability in the world today. It carries an enormous emotional and socioeconomic impact on patients, families, and health services. However, many strokes are preventable, and you could reduce your risk of having a stroke simply by being aware of, and controlling, certain lifestyle factors.

Did you know that 1 in 4 people will have a stroke in their lifetime? Knowing your risk factors for stroke may help you to prevent stroke.

We are running a study to compare two different ways of showing people their risk factors for stroke. We will test whether one method is better than the other to helping people change their stroke risk factors, which are things like diet, exercise and blood pressure.

**How is the study being funded?**

This is an investigator initiated study that is funded as part of a 5-year grant from the National Health and Medical Research Council (NHMRC).

**Why have I been invited to participate?**

You are eligible to take part in this study if you are aged between 35 and 75; have at least two risk factors for stroke; have a smartphone (e.g. iPhone or Samsung Galaxy).

You may not be able to participate if you do not have a smartphone; have a terminal illness; have had a stroke or other cardiovascular event, like a heart attack; are already in another study to manage your stroke risk factors; have a family member participating in this study; or have problems with memory or thinking.

**What will I be asked to do?**

Firstly, to confirm eligibility for the study, you will be asked to complete a short health and physical activity online questionnaire. This will ask for information about you and your health including your diet, level of physical activity, blood pressure, your weight and height, and daily tobacco intake. If you are eligible to participate in the study, you will then be asked to complete a short quiz at your face-to-face appointment to test whether you have any problems with memory or thinking. If you are found to have problems with memory and thinking on this quiz you will not be eligible to participate. We expect the online health questionnaire to take no more than 30 minutes. Your results of both the health questionnaire and memory quiz will determine your eligibility to participate in the study.

To continue with the study, if you agree to participate, you will be sent a link via e-mail to book an appointment for a health assessment and to complete some more detailed online questionnaires about your health, medical conditions, diet, exercise, quality of life and psychological wellbeing. You can complete the questions online in your own time, which will take about 30 minutes. All questions are voluntary, and you can choose whether to answer or not.

The face-to-face appointment will be in a central location and can be made at a time convenient to you. At the clinic, a research assistant will take measurements of your risk factors including height, weight, blood pressure, blood lipids and glucose level. We will also seek your permission to access your Medicare Benefits Schedule (MBS) and Pharmaceutical Benefits Scheme (PBS) data from Services Australia. Medicare collects information on your doctor visits and the associated costs, while the PBS collects information on the prescription medications you have filled at pharmacies. Services Australia supplies the data with your consent. This data then undergoes a process which is called data linkage.

is done by specially trained staff in secure facilities to protect your privacy. With your consent we provide your personal details securely that can then be linked to your use of health services including medications to help manage cardiovascular or metabolic diseases because these will help us to understand how the different ways of presenting the information about your risk factors may have changed health service use.

After the assessment you will be randomized to receive your risk factor information in one of two different ways. You cannot choose which group you are put into. You will be required to read the information we provide to you and follow links to information via your mobile phone. The minimum time required for each intervention is around 10 minutes for reading but you can choose how much time you spend reading the information. Depending on the group you are assigned to, you may receive some brief correspondence (e.g. 1 to 2 minutes reading time) from the study investigators about your risk factors over the next 6 months. You may receive also an additional short questionnaire online 1 week after the first assessment to follow-up about the summary you received. This will take less than 5 minutes to complete.

There will then be three follow-up appointments to complete over 12 months:

1. Online questionnaires at 3 months – 30 minutes
2. Face-to-face health assessment 6 months – 60 minutes
3. Online questionnaires at 12 months – 30 minutes

This will enable us to see if there has been any improvement over time.

You will also be asked to complete a brief satisfaction survey after the 6- and 12- month assessment. There is also the option to participate in a group or individual interview to further explore your experiences with the study.

We expect the total amount of time over the twelve-month period being around 3½ to 4 hours to participate in screening, baseline, 3, 6, and 12-month follow-up assessments.

**Are there any possible benefits from participation in this study?**

The main benefit for you is to learn more about your individual stroke risk factors. All the physical measurements will be done for free and you will receive a full profile of your risk factors for stroke. You can also be reimbursed for your time and travel costs if needed to a total value of $50 ($25 at the start and $25 at the end of the study).

Overall, information gained from this research has the potential to improve primary prevention of stroke, heart attack, dementia, diabetes mellitus and some types of cancer.

**Are there any possible risks from participation in this study?**

There may be some minor discomfort from the blood-test done via a finger-prick to measure blood lipids and glucose. We will also ask questions about your physical health and psychological wellbeing, which may make you feel uncomfortable. You can choose to not answer questions if they make you too uncomfortable. We will provide links to high quality information to help you if needed. Our data collectors are well-trained, and they will provide you with the ability to opt out of any questions or assessment and to withdraw from study at any time without the need to explain why. If you require further information about the study or the questions, you will be able to contact the researcher by email or phone. The contact details are at the beginning of this document.

**Your participation is voluntary**

Your participation in this study is completely voluntary and there will be no cost to you. If you do not want to take part in this study you do not have to. You should feel under no obligation to participate in this study. Choosing not to take part in this study will not affect your current and future medical care in any way.

**Your withdrawal from the study**

You are under no obligation to continue with the research study. You may change your mind at any time about participating in the research. People withdraw from studies for various reasons and you do not need to provide a reason.

You can withdraw from the study at any time by completing and signing the “Participant Withdrawal of Consent Form”. This form is provided at the end of this document and is to be completed by you and supplied to the research team if you choose to withdraw at a later date.

If you withdraw from the study, you will be able to choose whether the study will destroy or retain the information it has collected about you. You should only choose one of these options. Where both boxes are ticked in error or neither box is ticked, the study will destroy all information it has collected about you.

**What will happen to the information about me?**

Your information will be stored, retained and destroyed in accordance with relevant Australian and/or South Australian, Tasmania or Victorian, privacy and other relevant laws’. All information collected for this study will be stored in a secure database on a University of Tasmania server. This database is password-protected and only accessible to the researchers of this study. The study manager will control access and monitor database use. At the conclusion of the study all data will be non-identifiable. It will be stored on a University of Tasmania server for 15 years from the publication of results of this study. Data will be securely destroyed at the end of the 15 year period and done in such a way that the data cannot be recovered.

Any extension to these storage periods would only occur with ethical approval from the approving institution.

Any paper documents resulting from the study such as consent forms will be stored in accordance with the University of Tasmania’s data storage policy. Paper study documents will be stored in a locked draw in a restricted research facility. Only members of the research team who require this information will be given access.

In accordance with relevant Australian and/or South Australia, Tasmania or Victorian privacy and other relevant laws, you have the right to request access to your information collected and stored by the research team. You also have the right to request that any information with which you disagree be corrected. Please contact the study team member named at the end of this document if you would like to access your information.

**What if I change my mind during or after the study?**

You are free to withdraw without consequence before your 12-month follow-up assessment. There is a form to complete to withdraw from the study and you can choose to either keep your data in the study or have it destroyed. At up to 12 months after the start of the study it is easier for us to remove your data, after this time your data may have been included in the analysis and it will not be possible to remove it because it will have been de-identified and potentially included in publications.

**How will the results of the study be published?**

All published data from this study will be de-identified, without the possibility of re-identification of participant data at any stage. We will only use the information you provide to this study, including your MBS and PBS data, for the purpose of this study. The consent form allows you to choose if we can only use your data for this study or if we can also use it in other studies. This consent also relates to the data we collect from the Medicare Benefits Schedule (MBS) and Pharmaceutical Benefits Scheme (PBS). If the data is used in other studies, these will be approved by an ethics committee and your data will be de-identified to protect your privacy. We will provide you with a summary of the study findings by e-mail when it has finished.

**What if I have questions about this study?**

If you have any queries, concerns or issues with this study, please feel free to contact us:

Tasmanian coordinating centre:

Ass/Pro Seana Gall

South Australian contact:

Prof Timothy Kleinig

Victorian contact:

Prof Amanda Thrift

This study has been approved by the Tasmania Health and Medical Human Research Ethics Committee, [names of other committees here].

For matters relating to research at the site at which you are participating, the details of the local site complaints person are:

*Tasmania*
Executive Officer of the HREC (Tasmania) Network on (03) 6226 6254 or. The Executive Officer is the person nominated to receive complaints from research participants. You will need to quote H0023615.

*South Australia*
CALHN Research Office Manager, Ms Bernadette Swart, Ph (08) 7117 2209,.

*Victoria*

Monash University Research and Ethics Office, Ph (03) 9902 0132,

**How can I agree to be involved?**

If you are interested in participating in this study, please complete the initial questionnaire at: <https://redcap.utas.edu.au/surveys/?s=M7N8DHFE3J>

To be involved further we will ensure your informed consent to the study. There two aspects to your consent, the first being participation in the study as described above, and the second being that you authorise the study to access your complete Medicare Benefits Schedule (MBS) and Pharmaceutical Benefits Scheme (PBS) data as outlined in the consent form. This part of the consent form is sent securely to Services Australia who holds the MBS and PBS data confidentially.

If you have a privacy complaint in relation to the use of your MBS/PBS data you should contact the Office of the Australia Information Commissioner. You will be able to lodge a complaint with them.

Website: www.oaic.gov.au

Mail: GPO Box 5218, Sydney NSW 2001

If you have a privacy complaint in relation to the use of general study data you should contact the Privacy Commissioner in your relevant state. You will be able to lodge a complaint with them.

New South Wales

Website: www.ipc.nsw.gov.au

Mail: GPO Box 7011, Sydney NSW 2001

Queensland

Website: www.oic.qld.gov.au

Mail: PO Box 10143, Adelaide Street, Brisbane, Queensland 4000

Victoria

Website: www.ovic.vic.gov.au

Mail: PO Box 24274, Melbourne VIC 3001

**Thank you for your time**

**PARTICIPANT CONSENT FORM**

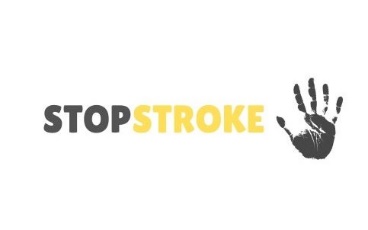

| Research team | Associate Professor Seana Gall*,* Menzies Institute for Medical Research, University of Tasmania   Professor Timothy Kleinig  Royal Adelaide Hospital   Professor Amanda Thrift  Monash University   Professor Dominique Cadilhac  Monash University  |
| --- | --- |

By signing below, I confirm that I have been provided with information and understood the information sheet and in particular:

- I understand that my involvement in this research will include answering questions about my health, medical conditions, diet, lifestyle, and psychological wellbeing. These questions will be answered at the commencement of the study, then again at 3, 6 and 12 months.
- I understand my involvement in this research will include attending two face-to-face interviews where physical measurements will be taken, including a blood test. The face-to-face interview will be at the commencement of the study and again at 6 months.
- I understand that participation may risk some discomfort arising from the blood test and answering questions about my physical and emotional wellbeing. I understand that if I do feel uncomfortable, I am not obliged to answer all the questions and may withdraw from the study at any time.
- Details of procedures and any risks have been explained to my satisfaction.
- I have been given the opportunity to ask questions and any questions that I have asked have been answered to my satisfaction.

I understand that all study data will be securely stored on the University of Tasmania premises for fifteen years from the publication of the study results, and will then be destroyed unless I give permission for it to be used to support other research in the future.

Please select at least one of the following

☐	I agree that my study data can be used for this specific project

☐	I agree that my de-identified study data can be shared and used for future research projects in the same general area of this research

☐	I agree that my de-identified study data can be shared and used for any future research

- I understand that the results of the study will be published so that I cannot be identified as a participant.
- I understand that my participation in this research is voluntary.
- I understand that I am free to withdraw at any time, without explanation or penalty.
- If I wish, I may request that any data I have supplied be withdrawn from the research until 12 months from this day.
- I agree to participate in the study.

| Name |
| --- |
| Signature |
| Date |

| **Statement by Researcher** | |
| --- | --- |
|  | I have explained the project and the implications of participation in it to this volunteer and I believe that the consent is informed and that he/she understands the implications of participation. |
| If the researcher has not had an opportunity to talk to participants prior to them participating, the following must be ticked. | |
|  | The participant has received the Information Sheet where my details have been provided so participants have had the opportunity to contact me prior to consenting to participate in this project. |

| Name |
| --- |
| Signature |
| Date |

### Supplementary Table 1. Definitions for scoring of the individual LS7 items to create overall LS7 scores

Each item is scored 2 for ideal, 1 for intermediate and 0 for poor according to scoring criteria in table below, adapted from American Heart Association recommendations.^2^ The total score for each participant is the sum of the 7 individual items, ranging from 14 (ideal cardiovascular health) to 0 (poor cardiovascular health). To summarise the level of cardiovascular health, the total score was also categorised into ‘high’ (scores 9-14), ‘intermediate’ (scores 6 to 8) and ‘low’ (scores 0 to 5).

|  |  | **Score** | | |
| --- | --- | --- | --- | --- |
| **Risk factor** | **Measure and scoring** | **2 = ideal** | **1 = intermediate** | **0 = poor** |
| Smoking status | Self-report questionnaire | Never smoker or quit smoking> 12 months ago | Former smoker ≤ 12 months | Current smoker |
| Body mass index | Measured height and weight | < 25 kg/m^2^ | 25–29.9 kg/m^2^ | ≥ 30 kg/m^2^ |
| Physical activity | International Physical Activity Questionnaire (IPAQ) short version  IPAQ categories for high and moderate physical activity used for LS7 categories of ‘ideal’ and ‘intermediate’ respectively | ≥ 3 days per week of vigorous physical activity for  ≥1500 MET mins/week; or 7 days per week of any activity for ≥3000 MET mins/week | ≥ 3 days per week of vigorous physical activity for ≥ 20 minutes per week; or ≥ 5 days per week of total activity for ≥ 600 MET minutes per week; or ≥ 5 days per week of moderate or walking activity for ≥ 30 minutes per week | Levels of physical activity not meeting high or moderate levels |
| Healthy diet score | Diet Quality Score (DQS)– brief food frequency questionnaire comprising fats, non-milk extrinsic sugars, vegetable, fruit and fish intakes with total score from 5 (poor diet) to 15 (good diet). | DQS >12 and ≤15 | DQS >8 and ≤12 | DQS ≥5 and ≤8 |
| Total cholesterol | Measured from point of care non-fasting test | < 5.18mmol/l | 5.18-6.18mmol/l or <5.18 mmol/L with medication | ≥ 6.18mmol/l |
| Blood pressure | Measured using Omron automated blood pressure monitor with standard protocol including sitting quietly for 5 minutes with average of 3 measures taken | SBP < 120 mmHg and DBP< 80 mmHg | SBP 120–139 mm Hg or DBP 80–89 mm Hg or  SBP < 120 mmHg and DBP< 80 mmHg with medication | SBP ≥ 140 mmHg or DBP ≥ 90 mmHg |
| Blood glucose | Measured from point of care non-fasting test | < 5.5mmol/l | ≥5.5 to≤6.94mmol/l or <5.5 mmol/L with medication | > 6.94 mmol/l |

MET – metabolic equivalent of task; SBP- systolic blood pressure; DBP- diastolic blood pressure

### Supplementary Table 2. Summary of assessments at baseline, 3, 6 and 12 months

| **Outcome measure** | **Baseline** | | **3 months** | **6 months** | | **12 months** |
| --- | --- | --- | --- | --- | --- | --- |
|  | **Online** | **Face to face** | **Online** | **Online** | **Face to face** | **Online** |
| Demographic and socioeconomic factors | x |  |  |  |  |  |
| Marital and employment status | x |  | x | x |  | x |
| Cognitive function (MoCA)^3^ |  | x |  | x |  | x |
| Primary outcome - Life’s Simple 7 |  | x |  |  | x |  |
| Diet quality score^4^ | x |  | x | x |  | x |
| Physical activity (IPAQ)^5^ | x |  | x | x |  | x |
| Smoking – current and historical | x |  | x | x |  | x |
| Non-fasting cholesterol – point of care test |  | x |  |  | x |  |
| Non-fasting glucose – point of care test |  | x |  |  | x |  |
| Measured height and weight for BMI |  | x |  |  | x |  |
| Blood pressure – automated monitor^6^ |  | x |  |  | x |  |
| Secondary outcomes | x |  |  | x |  | x |
| Psychological wellbeing (PHQ-9)^7^ | x |  | x | x |  | x |
| Stroke awareness (recognition of risk factors, knowledge of actions) | x |  | x | x |  | x |
| Quality of life (EQ-5D-5L)^8^ | x |  | x | x |  | x |
| Frequency of App use and Absolute 5-year risk of stroke (AUT Riskometer database) | x |  | x | x |  | x |
| Resource use questionnaire | x |  |  | x |  | x |
| Health costs: Data linkage – MBS/PBS (Australia); NMDS (New Zealand) |  |  |  |  |  | x |
| Participant satisfaction questionnaire |  |  |  | x |  | x |

MoCA; Montreal Cognitive Assessment, PHQ-9; Patient Health Questionnaire – 9, BP; Blood Pressure, BMI; Body Mass Index, HR; Heart Rate, IPAQ; International Physical Activity Questionnaire, AUT; Auckland University of Technology, CVD; Cardiovascular disease, MBS; Medicare Benefits Schedule, PBS; Pharmaceutical Benefits Scheme, NMDS; National Minimum, Dataset.

### Supplementary Table 3. Baseline LS7 items including missing data by intervention group

| **Item** | **Level** | **Usual care group**  **n (%)** | **Intervention group**  **n (%)** | **Total**  **n (%)** |
| --- | --- | --- | --- | --- |
| Physical activity | Poor | 124 (29%) | 147 (34%) | 271 (31%) |
|  | Intermediate | 181 (42%) | 157 (37%) | 338 (39%) |
|  | Ideal | 128 (30%) | 125 (29%) | 253 (29%) |
| Diet | Poor | 34 (8%) | 42 (10%) | 76 (9%) |
|  | Intermediate | 345 (80%) | 328 (76%) | 673 (78%) |
|  | Ideal | 54 (12%) | 59 (14%) | 113 (13%) |
| Smoking | Poor | 38 (9%) | 45 (10%) | 83 (10%) |
|  | Intermediate | 8 (2%) | 7 (2%) | 15 (2%) |
|  | Ideal | 387 (89%) | 377 (88%) | 764 (89%) |
| Cholesterol* | Poor | 34 (8%) | 46 (11%) | 80 (9%) |
|  | Intermediate | 165 (38%) | 190 (44%) | 355 (41%) |
|  | Ideal | 234 (54%) | 193 (45%) | 427 (50%) |
| Blood glucose | Poor | 79 (18%) | 68 (16%) | 147 (17%) |
|  | Intermediate | 86 (20%) | 102 (24%) | 188 (22%) |
|  | Ideal | 268 (62%) | 259 (60%) | 527 (61%) |
| Blood pressure | Poor | 176 (41%) | 197 (46%) | 373 (43%) |
|  | Intermediate | 209 (48%) | 183 (43%) | 392 (45%) |
|  | Ideal | 48 (11%) | 49 (11%) | 97 (11%) |
| Body mass index (BMI) | Poor | 214 (49%) | 226 (53%) | 440 (51%) |
|  | Intermediate | 152 (35%) | 142 (33%) | 294 (34%) |
|  | Ideal | 67 (15%) | 61 (14%) | 128 (15%) |
| Total LS7 score at baseline | Low (0-6) | 90 (21%) | 110 (26%) | 200 (23%) |
|  | Intermediate (7-8) | 149 (34%) | 155 (36%) | 304 (35%) |
|  | High (9-14) | 194 (45%) | 164 (38%) | 358 (42%) |

*χ^2^ test p<0.05 usual care versus intervention group

### Supplementary Table 4. Protocol violations including missing primary outcome data, loss to follow-up and other protocol violations related to study processes

| **Type of protocol violation** | **Level** | **Usual care group**  **N=433**  **n** | **Intervention group**  **N=429**  **n** |
| --- | --- | --- | --- |
| Missing at least one LS7 item at 6 months |  | 76 | 95 |
| Reason for missing LS7 items at follow-up | Withdrawn or lost to follow-up | 33 | 45 |
|  | No 6-month questionnaire data | 0 | 1 |
|  | No 6 month physical measurements | 22 | 21 |
|  | Error in physical activity item | 13 | 20 |
|  | Error in other item | 8 | 8 |
| Other protocol violations | Received e-mail about app – potential unblinding in usual care group | 70 |  |
|  | Research assistant potentially unblinded | 1 | 31 |
|  | Did not report downloading or using app at 6 months |  | 90 |
| Combined protocol violations |  | 113 | 153 |

### Supplementary Table 5. Differences in mean overall LS7 score difference from baseline to 6 months by pre-specified subgroups of gender, SES, country, and baseline LS7 group

|  |  | **Unadjusted model** | | | **Adjusted model*** | | |
| --- | --- | --- | --- | --- | --- | --- | --- |
|  |  | **Difference in mean LS7 score differences between groups*** | **(95% CI)** | **p-value** | **Difference in mean LS7 score differences between groups*** | **(95% CI)** | **p-value** |
| Sex | Male | -0.14 | -0.50, 0.23 | 0.467 | -0.13 | -0.49, 0.24 | 0.498 |
|  | Female | 0.12 | -0.16, 0.40 | 0.374 | 0.17 | -0.11, 0.45 | 0.228 |
| SES | 1 (Lowest) | -0.39 | -0.95, 0.17 | 0.171 | -0.36 | -0.92, 0.20 | 0.206 |
|  | 2 | 0.21 | -0.42, 0.85 | 0.489 | 0.18 | -0.46,0.82 | 0.559 |
|  | 3 | 0.15 | -0.33, 0.63 | 0.525 | 0.14 | -0.34, 0.63 | 0.550 |
|  | 4 | 0.01 | -0.42, 0.44 | 0.963 | 0.02 | -0.41, 0.45 | 0.915 |
|  | 5 (Highest) | 0.16 | -0.30, 0.63 | 0.468 | 0.20 | -0.27, 0.67 | 0.380 |
| Age category (years) | 35-45 | 0.05 | -0.51, 0.61 | 0.862 | 0.09 | -0.47, 0.66 | 0.749 |
|  | 46-55 | -0.08 | -0.54, 0.39 | 0.732 | -0.09 | -0.56, 0.37 | 0.682 |
|  | 56-65 | -0.03 | -0.44, 0.38 | 0.867 | 0.00 | -0.41, 0.40 | 0.981 |
|  | 66-75 | 0.19 | -0.20, 0.56 | 0.313 | 0.21 | -0.19,0.60 | 0.286 |
| Country | Australia | 0.08 | -0.22, 0.39 | 0.588 | 0.11 | -0.20,0.41 | 0.500 |
|  | NZ | -0.03 | -0.35, 0.29 | 0.857 | -0.00 | -0.33, 0.32 | 0.987 |
| Baseline LS7 group | High | 0.03 | -0.32, 0.37 | 0.863 | 0.03 | -0.31, 0.38 | 0.841 |
|  | Intermediate | 0.19 | -0.19, 0.57 | 0.315 | 0.22 | -0.17,0.60 | 0.255 |
|  | Low | -0.22 | -0.65, 0.22 | 0.324 | -0.14 | -0.58, 0.29 | 0.514 |

SES- All interactions between subgroups and intervention group were p>0.05. * Adjusted for age, sex, country and SES

### Supplementary Table 6. Per protocol analysis for mean within person differences in individual LS7 items and differences in the mean differences between usual care and interventions groups from baseline to 6 months

| **Item^1^** | **Adjusted**  **baseline**  **mean***  **(95% CI)** | **Mean difference over time in usual care group***  **(95% CI)** | **Mean difference over time in intervention group***  **(95% CI)** | **Difference in mean differences between treatment groups***  **(95% CI)** | **P-value for difference in differences** |
| --- | --- | --- | --- | --- | --- |
| BMI (kg/m^2^) | 30.22 (28.17, 32.27) | -0.09 (-0.26, 0.08) | -0.07 (-0.26, 0.11) | -0. 01 (-0.23, 0.26) | 0.92 |
| Physical activity (MET mins/week) | 1996.49 (1317.66, 2675.54) | 120.01 (-144.34, 384.36) | 514.25 (229.53,798.97) | 394.24 (27.80, 760.67) | 0.04 |
| Dietary quality score | 10.05 (9.65, 10.46) | 0.14 (-0.01, 0.30) | 0.29 (0.13, 0.46) | 0.15 (-0.06, 0.36) | 0.17 |
| Blood glucose (mmol/L) | 5.46 (4.95, 5.97) | 0.16 (-0.06, 0.38) | 0.11 (-0.13, 0.34) | -0.05 (-0.34, 0.24) | 0.74 |
| Systolic blood pressure (mmHg) | 128.93 (124.39, 133.46) | -3.23 (-4.71, -1.75) | -2.28 (-3.86, -0.71) | 0.95 (-1.11, 3.00) | 0.37 |
| Total cholesterol (mmol/L) | 4.41 (4.32, 4.50) | 0.04 (-0.07, 0.15) | 0.08 (-0.04, 0.20) | 0.04 (-0.11, 0.19) | 0.64 |
| Proportion of current smokers | 0.108 (0.099, 0.116) | -0.003 (-0.010, 0.005) | -0.005 (-0.009, 0.003) | -0.003 (-0.013,0.008) | 0.60 |

^1^ Using only the continuous component of each item and not the qualifying factors that are included in the total LS7 calculation. For example, systolic blood pressure as used for the item analysis is not adjusted according to use of blood pressure medication.

MET - * Adjusted for age, sex, country and SES. Baseline mean is presented at reference levels: age category = 35-45 years, country= Australia, SES quintile= 3 (middle) and sex= male.

Results estimated using mixed models

### Supplementary material 7 - References

1.	Parmar P, Krishnamurthi R, Ikram MA, Hofman A, Mirza SS, Varakin Y, et al. The S troke R iskometer TM A pp: Validation of a data collection tool and stroke risk predictor. *Int J Stroke* 2015; 10: 231-244.

2.	Lloyd-Jones DM, Hong Y, Labarthe D, Mozaffarian D, Appel LJ, Van Horn L, et al. Defining and setting national goals for cardiovascular health promotion and disease reduction: the American Heart Association's strategic Impact Goal through 2020 and beyond. *Circulation* 2010; 121: 586-613.

3.	Nasreddine ZS, Phillips NA, Bédirian V, Charbonneau S, Whitehead V, Collin I, et al. The Montreal Cognitive Assessment, MoCA: A brief screening tool for mild cognitive impairment. *J Am Geriatr Soc* 2005; 53: 695-699.

4.	Cleghorn CL, Harrison RA, Ransley JK, Wilkinson S, Thomas J and Cade JE. Can a dietary quality score derived from a short-form FFQ assess dietary quality in UK adult population surveys? *Public Health Nutr* 2016; 19: 2915-2923.

5.	Booth M. Assessment of physical activity: an international perspective. *Res Q Exerc Sport* 2000; 71: S114-120.

6.	Mancia G, Fagard R, Narkiewicz K, Redon J, Zanchetti A, Bohm M, et al. 2013 ESH/ESC Practice Guidelines for the Management of Arterial Hypertension. *Blood Press* 2014; 23: 3-16.

7.	Kroenke K, Spitzer RL and Williams JB. The PHQ-9: validity of a brief depression severity measure. *J Gen Intern Med* 2001; 16: 606-613.

8.	Golicki D, Niewada M, Buczek J, Karlińska A, Kobayashi A, Janssen M, et al. Validity of EQ-5D-5L in stroke. *Qual Life Res* 2015; 24: 845-850.
